# Diagnostic Accuracy of an Immunoassay Using Avidity-Enhanced Polymeric Peptides for SARS-CoV-2 Antibody Detection

**DOI:** 10.64898/2026.02.26.26343835

**Authors:** Brian Andrich L. Pollo, Danica S. Ching, Maria Isabel C. Idolor, Ruby Anne N. King, Fresthel Monica M. Climacosa, Salvador Eugenio C. Caoili

## Abstract

**Background:** There is a need for synthetic peptide-based serologic assays that exploit avidity to replace whole antigens while enabling low-cost diagnostics in resource-limited settings.

**Objective:** To evaluate the diagnostic accuracy of a polymeric peptide-based ELISA leveraging avidity to enhance signal.

**Method:** A 15-member SARS-CoV-2 peptide library corresponding to multiple epitope clusters and proteins was screened by indirect ELISA using pooled sera from RT-PCR-confirmed COVID-19 patients to identify peptides with possible diagnostic utility. The identified lead candidate, S559, possessed terminal cysteine-substitution to allow disulfide polymerization, and the resulting avidity gain was evaluated by comparing the apparent dissociation constant (K_D_^app^) before and after depolymerization with N-acetylcysteine. The performance of an optimized ELISA using S559 was evaluated on 1,222 prospectively collected COVID-19 serum samples and 218 biobanked pre-COVID control serum samples.

**Results:** Polymeric S559 with a K_D_^app^ of 29.26 nM^-1^was demonstrated to have a 218% avidity gain relative to the completely depolymerized form. At pre-defined thresholds, the optimized S559 ELISA has a sensitivity and specificity of 83.39% (95%CI: 81.18% and 85.43%) and 96.79% (95%CI: 93.50% and 98.70%), respectively. At *post hoc* thresholds determined by Youden index, sensitivity and specificity reached 95.01 (95% CI: 93.63% - 96.16%) and 100.00% (95% CI: 98.32% - 100.00%), respectively.

**Conclusion:** Homomultivalent epitope presentation using polymeric S559 allows a highly specific immunoassay using human sera that may have important value in detecting antibodies, whether for diagnosing infection, confirming vaccination status or conducting surveillance.

## 1. INTRODUCTION

Peptides have been used in serologic assays (*e*.*g*., enzyme-linked immunosorbent assay or ELISA) for the detection of antibodies [1,2], which in turn are correlates of infectious diseases [3,4]. These assays typically employ monovalent antigen formats that enable singular binding events. However, such monovalent interactions may exhibit limited sensitivity, especially when detecting low-affinity or low-titer antibodies [5–8].

This is particularly important during the early stages of a disease or in cases of mild or asymptomatic infections, where the antibody response can be weak or inconsistent [9–12]. The diagnostics for severe acute respiratory syndrome coronavirus 2 (SARS-CoV-2), the virus responsible for coronavirus disease 19 (COVID-19), are notably impacted, as the humoral immune response can vary widely and tends to decrease over time [13–16].

To address such challenges with peptide antigens, multimeric or polymeric arrangements have been explored. These designs aim to replicate the repetitive and clustered structures of antigens that are naturally found on the surfaces of pathogenic particles, such as the trimeric spike glycoproteins of SARS-CoV-2. For instance, peptides representing multiple epitopes have been combined on scaffolds, conjugates or carriers [13–17]. Simpler, economical approaches have been employed as well, using cysteine-substituted peptides [18]. By presenting antigens in a multivalent format, these assays can enhance avidity, allowing multiple B-cell receptors or antibodies to engage simultaneously [19,20]. This boosts overall binding strength, surpassing what monovalent formats can achieve [21,22]. Such improvements in avidity can lead to better assay sensitivity and a more favorable signal-to-noise ratio, especially for antibodies that have weak monovalent affinities [23].

Despite these advancements, there remains an unmet need in COVID-19 for well-defined peptide antigens that are specifically designed to leverage avidity effects in immunoassay platforms. Ideally, these peptides would present conformationally relevant epitopes of SARS-CoV-2 in a multivalent setup, thus enhancing detection sensitivity.

The present study aims to design and evaluate a multimeric peptide-based ELISA to enhance detection sensitivity via avidity gain. Specifically, we employed disulfide-linked peptides representing SARS-CoV-2 B-cell epitopes. The diagnostic accuracy of the developed assay was then assessed using prospectively collected clinical samples for RT-PCR-confirmed hospitalized COVID-19 patients.

The rationale for this study is the early COVID-19 pandemic experience in settings such as in the Philippines, where molecular diagnostic capacity was limited, and rapid antibody tests were deemed unreliable [24]. In this context, a peptide-based serological platform could have provided a more accessible alternative to molecular and whole protein methods. Beyond SARS-CoV-2, these principles may inform the design of peptide-based diagnostics for emerging pathogens.

## 2. MATERIALS AND METHODS

This manuscript was prepared according to the STARD guidelines for reporting diagnostic accuracy studies [29]. Ethics approval was obtained from the University of the Philippines Manila Research Ethics Board (Code 2022-0492-01). The full study protocol may be accessed via the Health Research and Development Information Network (HERDIN) under the registry number PHRR230601-005772. All authors had access to the study data and reviewed and approved the final manuscript.

### Peptide design

A SARS-CoV-2 synthetic peptide library was designed, synthesized, and assembled. Antigenic peptide design was carried out by systematically querying reported B-cell epitopes. The Immune Epitope Database was accessed on February 26, 2021. A search was performed on linear peptide B-cell epitopes with positive assay results originating from SARS-CoV-2 [organism ID:2697049]. Obtained epitopes were subjected to cluster analysis using the built-in function of the said database. Structure-function relationships were analyzed by retrieving three-dimensional structures from PDB and UniProt, and visualizing using PyMol. From these clusters, sequences were designed based on the following criteria (1) predicted intrinsic disorder, (2) length ≥12 amino acids and ≤20 amino acids[26], (3) absence of residues that may interfere with polymerization steps (*e*.*g*., C, M, W, K), and (4) <50% hydrophobicity. Sequence homology analysis was performed using BLASTp (NCBI), taking care to avoid nonspecific cross-reaction with non-SARS-CoV-2 epitopes. Designed peptide core sequences were subsequently modified by substitution of cysteine residues at N- and C-termini to facilitate polymerization via disulfide bond formation.

### Peptide synthesis and polymerization

Antigenic peptide synthesis was outsourced to GenScript (Singapore), a commercial provider. The peptides were synthesized using solid-phase chemistry. The peptides were then each analyzed by high-performance liquid chromatography (HPLC) on an Inertsil ODS-3 4.6 x 250 mm column using a mobile phase consisting of 0.065% trifluoroacetic acid (TFA) in 100% water (v/v) for solvent A and 0.05% TFA in 100% acetonitrile (v/v) for solvent B with a total flow of 1 ml per minute and time program set with 5% solvent B initially and 65% solvent B at 15– 25 minutes, with continuous UV absorbance monitoring of the eluate at 220 nm. Peptide polymerization was carried out under mild oxidizing conditions. DMSO was added up to a final concentration of 50% (v/v), then the resulting mixture was incubated for 24 hours at 25°C.

### Measurement of Avidity Gain by Depolymerization using Compositionally Matched Oxidation-Reduction with N-acetylcysteine (CORN)

The identified lead candidate peptide from screening was subjected to depolymerization to determine the avidity gain contributed by the polymeric structure. The depolymerized peptide was then subjected to derivatization with trinitrobenzene sulfonate (TNBS) and ELISA using anti-TNP as primary antibody. The apparent dissociation constant (K_D_^app^) was calculated for the polymeric peptide and the depolymerized form. Depolymerization was carried out using N-acetylcysteine (Nac), a reducing agent capable of breaking disulfide linkages. Aliquots of polymeric peptide were incubated with various concentrations of Nac for 30 minutes at 37°C. Prior to coating, sufficient DMSO was added to a final concentration of 20% to compositionally match the test solutions with the negative control. As a negative control, oxidized Nac (oNac) was prepared by heating Nac in the presence of 20% DMSO for 1 hour at 90°C. The complete degradation of Nac was confirmed using 5,5’-dithio-bis-(2-nitrobenzoic acid) as well as organoleptically with the evolution of the maize-like odor of dimethylsulfide. The said oNac was then incubated with aliquots of the polymeric peptide.

### Sulfitolysis

As an alternative depolymerization procedure to CORN, peptides were incubated with various concentrations of Na_2_SO_3_, ranging from 38:1 to 150:1, at 90°C for 30 minutes. As a negative control, heat-quenched sulfite was prepared by heating at 90°C for 2.5 hours.

### Indirect ELISA

Various concentrations of the peptide-Nac/oNac mixtures were diluted in coating buffer (carbonate–bicarbonate buffer, pH□9.6) for a final concentration of 10 nM peptide. High-binding 96□well microtiter plates were coated with the resulting mixtures. Each well received 100μL of the coating solution. The plates were incubated at 37°C for 1 hour. Following coating, wells were washed three times with PBS containing 0.05% Tween-20 (PBST). Derivatization was carried out by adding 100 μL of 10nM TNBS solution and incubating at 37°C for 15 minutes. Unreacted TNBS was quenched by adding 100 μL of 20 nM glycine and incubating at 37°C for 10 minutes. Wells were then washed three times with PBST. Unless otherwise specified, indirect ELISAs were performed using the following conditions for blocking, antibody addition, substrate development, and stopping. Non-specific binding sites were blocked by adding 200 μL 2% skimmed milk in PBST to each well and incubating at 37°C for 30 minutes. Wells were then washed three times with PBST. After washing for three times, 100 μL of 2-fold serial dilutions (starting concentration: 66.67 nM) of anti-TNP antibody was added per well and incubated for one hour at room temperature. After subsequent washing, 100 μL protein A peroxidase conjugate at 1:8000 was added to each well. After washing, 100 μL of chromogenic substrate solution was prepared from 1 mg 3,3′,5,5′-tetramethylbenzidine in phosphate citrate buffer (pH 5.0) and 2 μL 30% hydrogen peroxide. The plate was incubated at room temperature for 15 minutes to allow color development. The reaction was stopped with 100 μL 0.1M H_2_SO_4_. Absorbance values were then measured at 450 nm. Absorbance values were plotted against linearized antibody concentration to compute for K_D_^app^ [27–29]. The K_D_^app^ was calculated from the slope of the resulting linear plot.

### Peptide Library Screening

ELISA was performed using pooled sera from 20 patients with RT-PCR-confirmed COVID-19. Pooled sera from 20 healthy donors served as negative pre-COVID-19 controls. Fresh frozen plasma were sourced from the University of the Philippines – Philippine General Hospital Blood Bank, then recalcified using 25mM CaCl_2_. Indirect ELISA was performed to screen said sera, using 100μL of 20μg/mL peptide in coating buffer for coating, and 100μL pooled sera diluted to 1:100 as primary antibody. The rest of the steps such as blocking, washing, adding substrate, and stopping were performed as above.

### Immunoassay Optimization

Assays using the selected candidate peptides were subjected to further optimization. To confirm immobilization of the peptides on polystyrene plates, ELISA was performed using TNP-labelled peptides coated onto high-binding microtiter plates and detected with anti-TNP antibody. Indirect ELISA was performed as described in avidity gain determination, without the depolymerization step. Optimal antigen concentration was determined using pooled COVID-19 sera (*n* = 20), with 20μg/mL selected as the coating concentration for subsequent testing on clinical samples. The primary antibody used was 100μL pooled sera diluted to 1:100. Assay performance metrics were assessed across a serial geometric dilution of human serum (1:100 to 1:6400). For antigen concentration optimization and assay performance evaluation, indirect ELISA was performed as previously described, without the depolymerization and derivatization steps.

### Participants

To establish diagnostic performance, indirect ELISA was performed on prospectively collected clinical samples. All enrolled patients were (1) hospitalized at Philippine General Hospital, a 1,335-bed level III hospital, between October 2020 to February 2021, prior to vaccine availability in the country, (2) at least 18 years old and (3) with RT-PCR-confirmed COVID-19 from nasopharyngeal swab. Convenience sampling was utilized, with potentially eligible participants being referred by their physician-in-charge.

### Testing methods

RT-PCR testing was performed by hospital laboratory personnel, following standard operating procedures using a cycle threshold of 40 or less. Clinical data was available to medical technologists and pathologists for interpretation. Collection of serum samples coincided with routine blood extraction in the clinical setting on days 1, 7, and 14 of hospitalization. Day 1 of hospitalization coincides with day of enrollment. Disease severity was classified by their attending clinician according to their presentation on admission: mild (no pneumonia), moderate (with pneumonia), severe (greater lung involvement; requirement for supplemental oxygen), or critical (requirement for mechanical ventilation or signs of septic shock) [34]. Control serum samples from healthy blood donors obtained prior to the emergence of SARS-CoV-2 in December 2019 were used as negative controls for all assays. All serum samples were heat-inactivated at 56°C for 30 min and stored at −80°C before use. For the peptide ELISA, the predefined threshold was set at mean + 3SD of the no peptide controls. Research personnel performing ELISA were blinded to the disease status of the patient samples they were handling throughout the testing process.

### Diagnostic accuracy analysis

Measures of diagnostic accuracy were calculated using MedCalc Statistical Software version 23.0.8. The *post hoc* cutoff was calculated based on Youden J [35]. Measures were both calculated based on the prespecified threshold as well as the exploratory cutoff. There were no indeterminate test results and no missing data. Sample size calculation was performed using PASS 2020 power analysis and sample size software, with estimated prevalence of 42.3% and a null hypothesis that the ELISA will yield a binary result [36,37].

## 3. RESULTS

### Peptide Library Design, Synthesis and Screening

A peptide library comprising 15 unique sequences was designed, corresponding to 15 epitope clusters and 7 SARS-CoV-2 proteins, both structural and nonstructural (**Table 1**). The epitope maps illustrating the relative location of each epitope cluster are provided in **supplementary figure 1**. For the antigenic peptide S559, a single dominant peak was observed exhibiting an elution time of 19.550 minutes and comprising 46.512% of the integrated peak area (**supplementary figure 2A**). Mass spectrometry was performed on the said peak, yielding a mass-to-charge ratio (m/z) spectrum that showed dominant ions at m/z 1039.0 [M+2H]2+, as depicted in **supplementary figure 2B**. Analysis of said mass spectrum gave an estimated mass of 2076.32 Da, which closely approximates the theoretical mass of 2076.0 Da calculated for S559. The lyophilized peptides were reconstituted in sterile water then aliquoted and stored in −20°C until use. The polydisperse character of the polymeric peptide was demonstrated using SDS-PAGE under non-reducing conditions as shown in lane 2 of the Coomassie-stained gel of **supplementary figure 3A**. Densitometric analysis (**supplementary figure 3B)** revealed polymeric products with molecular weight as high as 50kDa, corresponding to 25-mers.

**Table 1.**
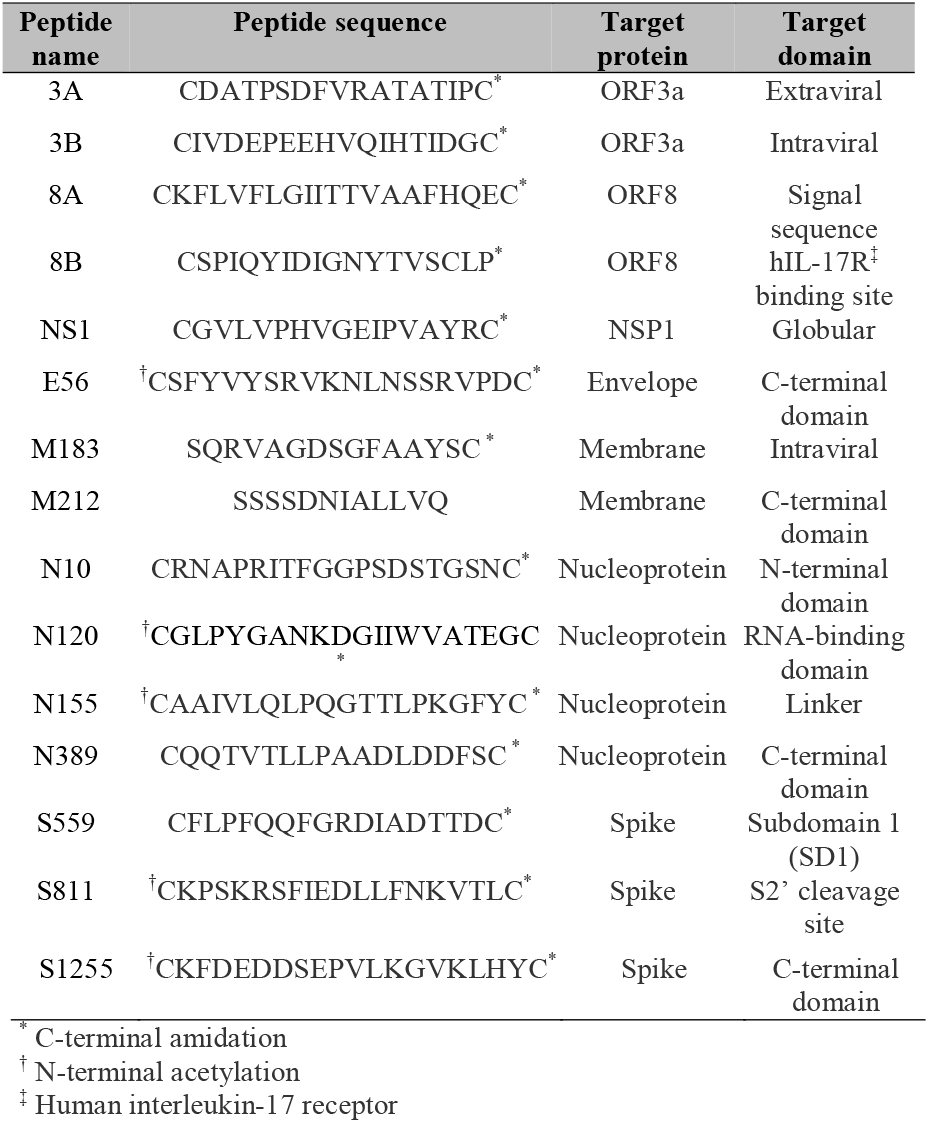
Sequences and targets of peptide library members.

### Avidity Gain Determination

Nac at a molar ratio of 100:1 relative to the peptide was able to facilitate complete depolymerization under the test conditions as shown in lane 3 Nac in **supplementary figure 3A**. The degree of depolymerization was comparable to that achieved by sulfitolysis at 38:1 molar ratio of sulfite to peptide. The estimated K_D_ ^app^ values were calculated from the linearized data of **supplementary figure 3C**, as shown in **Figure 2**. The polymeric peptide was found to have a K_D_ ^app^ of 29.26 nM^-1^, while the peptide depolymerized with 100:1 molar excess of N-acetylcysteine to peptide was found to have a K_D_ ^app^ of 63.76 nM-1. This corresponds to a 218% avidity gain obtained with polymeric S559.

**Figure 1.** Peptide library screening revealed lead peptides. ELISA was performed on pooled sera from 20 COVID-19 patients against pre-pandemic controls, showing relative reactivity of each peptide; data represent mean ± SD (*n* = 3); One-way ANOVA with *post hoc* Dunnett’s test vs. (-) ctrl (no peptide); * p ≤ 0.05, ** p ≤ 0.01, **** p ≤ 0.0001.

**Figure 2.**
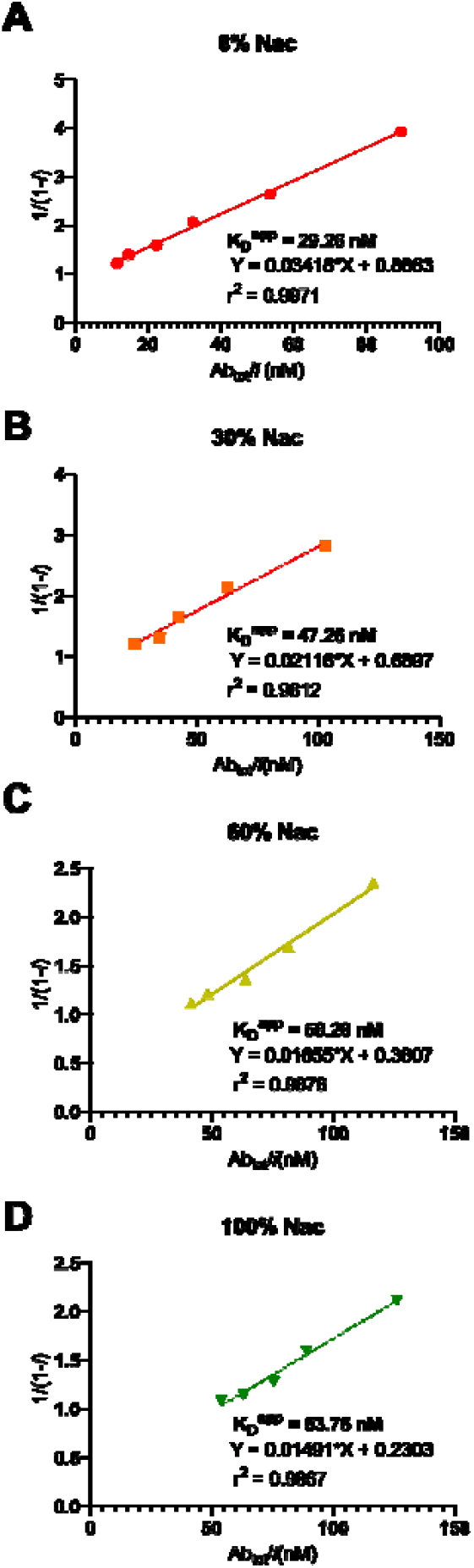
Linearized ELISA binding data demonstrating avidity gain of polymeric S559. Trinitrophenylated S559 was depolymerized with varying concentrations of Nac, then detected with anti-TNP antibody (αTNP). The resulting curves were used to estimate the apparent dissociation constants (K_D_^app^); 30% Nac: 300:70:1 molar ratio N-acetylcysteine : oxidized N-acetylcysteine : peptide; Ab_tot_: total antibody concentration; *i*: normalized OD450 reading.

### ELISA Optimization

Peptide library screening (**Figure 1**), identified S559 as the top candidate based on the COVID-19/pre-COVID-19 ratio. Antibodies against S811 and S1255, other peptides designed from spike, were also significantly increased in patients relative to controls. Antibodies against 3A, despite not being statistically significantly decreased, appears to be a negative predictor of COVID-19. To facilitate comparison, M212, N10, and E1 were further selected as comparator peptides representing their respective SARS-CoV-2 structural proteins. S559 exhibited a strong signal indicating successful and consistent plate immobilization (**Supplementary figure 4A**). Optimal antigen concentration was determined using pooled COVID-19 sera. Indirect ELISA was performed as described in avidity gain determination, without the depolymerization and derivatization steps. The primary antibody used was 100μL pooled sera diluted to 1:100. The signal plateaus at 20 µg/mL, which was subsequently used in all diagnostic ELISAs to balance signal strength and reagent cost (**Supplementary figure 4B**).

Assay performance metrics were assessed across a serial geometric dilution of human serum (1:100 to 1:6400) (**supplementary figure 4C**). The limit of detection corresponded to a serum dilution of 1:6400. The limit of quantitation was determined to be at a dilution of 1:1600. Intraassay coefficients of variation (CV%) were below 15% across the dynamic range, with CV% values ranging between 1.87% and 4.40%. A 1:100 dilution of sera was used for subsequent diagnostic assays. At this dilution, the signal-to-noise ratio was moderate at 5.82. Taken together with the low intraassay CV% values, this signal-to-noise ratio is consistent with a signal that is low in amplitude but consistently repeatable. The predetermined threshold was calculated as 0.057 absorbance units (AU) at 450nm.

### Diagnostic Accuracy Determination

A total of 1,222 prospectively collected serum samples were obtained from 544 hospitalized patients. Complete samples (baseline day7, and day14) were collected for 188 participants. For the rest of the participants, only 1 or 2 samples were collected because the patients were lost to various reasons (*e*.*g*., died or discharged from the hospital). **Table 2** shows the demographic and clinical characteristics of the final included participants. Among the samples tested, none were excluded for subsequent analyses. Results were compared against 218 pre-COVID-19 controls. At pre-defined thresholds, the optimized S559 ELISA has a sensitivity and specificity of 83.39% (95%CI: 81.18% and 85.43%) and 96.79% (95%CI: 93.50% and 98.70%), respectively. A cross-tabulation of ELISA results against RT-PCR results are depicted in **supplementary figure 5**, while a plot of individual sample results is shown in **supplementary figure 6**. Receiver operating characteristic (ROC) curve analysis of the assay data (**Figure 3**) produced an area under the ROC (AUROC) of 0.966 for polymeric S559. Under identical assay conditions, the two comparator antigens, M212 and N10, merely produced AUROC values of 0.739 and 0.583, respectively. The optimal cutoff OD450 value for the S559 assay was determined by Youden index to be 0.098AU. Sensitivity and specificity at this cutoff were 95.01 (95% CI: 93.63% - 96.16%) and 100.00% (95% CI: 98.32% - 100.00%), respectively. At a prevalence of 84.86%, the accuracy, positive predictive value, and negative predictive value are 95.76%, 100.00%, and 78.14% respectively. These findings are consistent with a sensitive and highly specific synthetic peptide-based diagnostic assay for COVID-19.

**Figure 3.**
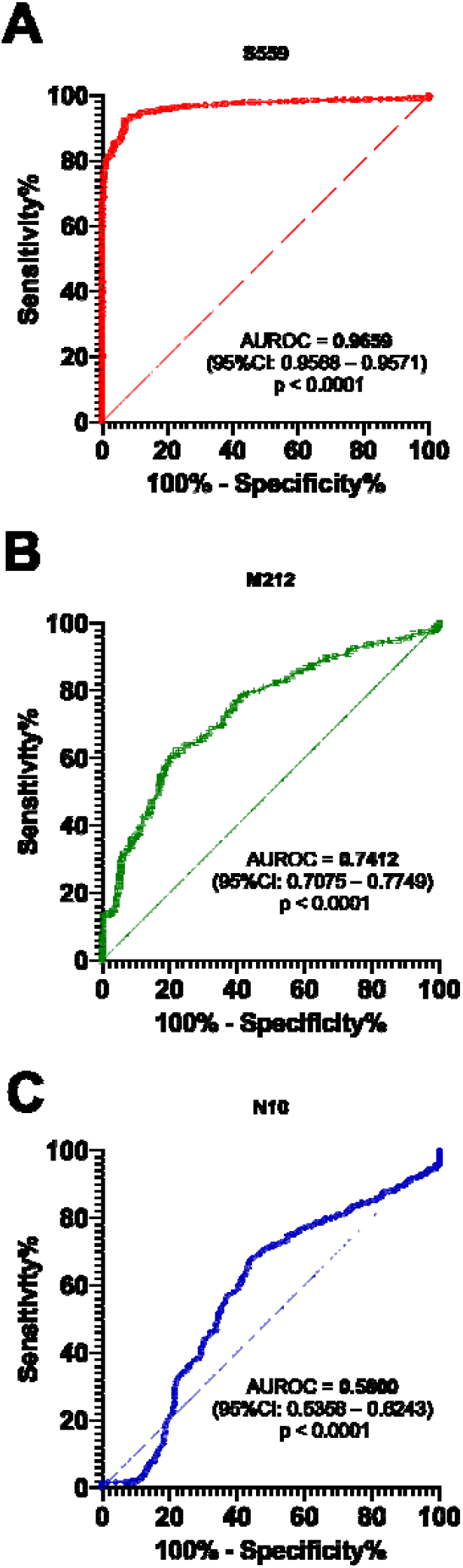
ROC curves demonstrating diagnostic performance of polymeric S559. Curves were generated using th**e O**D450nm values obtained from testing 1,222 samples and 218 controls. ROC: receiver operating characteristics; AUROC: area under the ROC curve.

**Table 2.** Baseline demographic and clinical characteristics of 544 participants yielding 1222 clinical case samples.

| Characteristic | Value |
| --- | --- |
| Sex* |  |
| Male | 334 (61) |
| Female | 210 (39) |
| Age | <sup>†</sup>54 ± 17<br><sup>‡</sup>71 |
| Disease severity* |  |
| Asymptomatic | 45 (8) |
| Mild | 47 (9) |
| Moderate | 119 (22) |
| Severe | 242 (44) |
| Critical | 91 (17) |
| Outcome* |  |
| Discharged | 414 (76) |
| Death | 84 (15) |
| Others<sup>§</sup> | 46 (9) |
| Sample contribution* |  |
| 1 sample<sup>†</sup> | 54 (10) |
| 2 samples<sup>**</sup> | 302 (55) |
| 3 samples<sup>††</sup> | 188 (35) |

\* no. (%) <sup>†</sup> mean ± SD (years) <sup>‡</sup> range (years) <sup>§</sup> e.g., absconded, transferred to hospital of choice <sup>†</sup> upon enrollment <sup>\*\*</sup> upon enrollment, day 7 <sup>††</sup> upon enrollment, day 7, day 14

## 4. DISCUSSION

This study aimed to develop an avidity-enhanced peptide-based diagnostic assay for COVID-19, focusing specifically on (1) identifying immunoreactive polymeric peptides designed from SARS-CoV-2, (2) demonstrating avidity gain with the candidate polymeric peptides, and (3) evaluating diagnostic accuracy of the resulting immunoassay.

By screening a library of rationally designed peptides, we identified several antigenic peptides. The pattern of immunoreactivity is consistent with known considerations in peptide design. Particularly, the high-reactivity peptides were based on surface-exposed domains of the spike protein. This is expected given the sheer number of copies of this protein on the viral surface, as well as its functional role in viral entry [38]. Interestingly, M212, despite its internal location, was relatively increased in COVID-19 cases as well. Such an epitope may function as decoy, eliciting an antibody response that does not confer protection [39]. The epitope represented by 3A appears to be a previously unreported inverse predictor. We hypothesize that antibodies against inverse predictors may be correlated with low disease severity, non-neutralizing response, or unrelated immune response. This is consistent with prior work in SARS-CoV-2 epitope profiling, wherein the cross-reactivity was observed between peptides from SARS-CoV-2 spike and nucleocapsid proteins and seasonal coronaviruses HCoV-OC43 and 229E [40,41]. Peptides from internally located nonstructural proteins have been observed to be reactive to pre-pandemic sera as well [42]. In this study, it should be noted that selected sequences were prescreened for sequence homology, thus potentially addressing the risk of cross-reactivity with seasonal coronaviruses.

However, these interpretations are limited by the inherent screening variability, given that peptides were coated at a uniform concentration. Intrinsic peptide-dependent properties such as hydrophobicity, charge, and folding propensity could influence coating efficiency on solid supports [43,44]. Thus, these differences introduce variability unrelated to antibody affinity or epitope accessibility, suggesting that signal magnitude is not a strict ranking of immunodominance or specificity [45]. Nevertheless, the observed pattern of immunogenicity provides clues to the future design of other peptides, highlighting the importance of utilizing epitopes located in structurally accessible and functionally important regions. Also, future work identifying diagnostically valuable epitopes must be careful of decoy regions and inverse predictors.

Yet another central finding in this study is the avidity gain observed with S559, a simple, self-polymerizing peptide, working as an antigen even in the absence of separate macromolecular scaffolds. Such an avidity gain is consistent with our hypothesis that homomultivalent antigenic presentation could considerably improve antibody binding, even in the absence of a scaffold or carrier. While most serological assays emphasize affinity, avidity reflects strength of multivalent binding. Traditional methods of assessing avidity in ELISA use chaotropic agents such as urea or sodium thiocyanate, agents which elute the antibody from the antigen by disrupting noncovalent interactions. However, these introduce the risk of eluting the adsorbed antigen from the plate, leading to misleading results [46–48]. In contrast, our CORN approach utilizing controlled disulfide breakage and depolymerization is a simpler strategy in estimating avidity contributions. A similar approach utilizing the reversibility of disulfide chemistry has been utilized before in larger enzymes [49,50]. While the accuracy of our dissociation constant estimate may yet be constrained by coating competition between the peptide and Nac, by pairing this specific depolymerization with our linearization strategy, we could estimate the dissociation constant without dependence on precise determination of coated antigen concentration. Further, the use of sulfur-based reducing agents (*e*.*g*., dithiothreitol, glutathione) relatively preserves native peptide conformation, unlike the ionic perturbation required for the nonspecific elution of chaotrope ELISAs. Future work may consider incorporating this approach into standardized diagnostic validation workflows, particularly for rationally designed minimal antigens such as disulfide-linked polymeric peptides.

We found that the developed S559 immunoassay was sensitive and highly specific. While the sensitivity falls short of the minimum >98% sensitivity recommended by the Philippine Department of Health and the World Health Organization [51], the excellent performance of S559 relative to the comparator peptide antigens is a step towards the development of peptide diagnostics that could match whole protein methods. Moreover, a synthetic peptide-based assay having a diagnostic performance rivaling that of full-length proteins is notable in itself [52]. Whole proteins, by virtue of conformational complexity as well as epitope diversity, typically yield higher sensitivity simply due to a broader range of antibody targets. Our findings suggest that a synthetic peptide representing a narrow linear fragment, specifically selected from rational design, can recapitulate meaningful diagnostic signals. While this minimalist, epitope focused peptide design offers advantages in cost, stability, and reproducibility, such an approach also carries the risk of diagnostic antigenic escape.

Another key observation is that the assay retained its sensitivity even in samples collected early in the disease course and from patients with mild or asymptomatic presentation. We speculate that the high sensitivity is a result of the avidity gain conferred by the homomultivalent structure of the polymeric peptide. This relatively excellent diagnostic performance, even among asymptomatic individuals, may be attributable to the avidity of the antigen, thereby facilitating stable binding even to early, low-affinity antibodies that might otherwise escape detection in conventional monovalent assays. The caveat being that the samples in this study may be underrepresented in terms of asymptomatic and mild cases.

This study is limited by its use of a hospital-based convenience cohort, leading to the overrepresentation of severe cases. Other limitations include variability in severity and day of illness. Stratification by these factors did not reveal any statistically significant difference in average signal intensity. The day of hospitalization does not necessarily reflect day of illness. Clinical severity was classified at the time of hospital admission and may not reflect subsequent disease progression or immune dynamics. Nonetheless, good performance metrics observed despite the variability in sampling times and disease severity suggests robustness under real-world clinical conditions.

Future studies should explore various peptide combinations and longitudinal seroconversion patterns beyond the 14-day window. Nonetheless, the current findings offer encouraging evidence that targeted peptide antigens can serve as diagnostic agents. It is conceivable that this immunoassay, combined in parallel or in series with other tests, could lead to improved sensitivity and specificity respectively.

## CONCLUSION

In this study, we developed a synthetic peptide-based ELISA designed to leverage avidity effects through disulfide polymerization of SARS-CoV-2 epitope sequences. The assay demonstrated promising diagnostic accuracy, including in samples from asymptomatic and mildly symptomatic individuals. These findings highlight the potential of simple homomultivalent peptide systems to enhance detection sensitivity by stabilizing low-affinity antibody interactions.

While limited by its single-center sample collection and convenience sampling strategy, the study illustrates the feasibility of using SARS-CoV-2 peptides in serological testing. Future work ought to assess performance in larger, more diverse populations (including vaccinated individuals, and those who have had prior infection) to assess generalizability.

## Data Availability

All data produced are available online at Pollo, Brian Andrich, 2026, "Replication Data for: Diagnostic Accuracy of an Immunoassay Using Avidity-Enhanced Polymeric Peptides for SARS-CoV-2 Antibody Detection", Harvard Dataverse

https://doi.org/10.7910/DVN/6WO6TG

## ACKNOWLEDGEMENTS

The authors would like to express their appreciation to the Department of Science and Technology Philippine Council for Health Research and Development (DOST-PCHRD) for providing support through scholarships and dissertation grants. The Section of Blood Banking and Transfusion Medicine, Department of Laboratories, University of the Philippines - Philippine General Hospital (UP-PGH) provided blood samples for testing. Sample collection was conducted as part of the “Collection and Archiving of Patient Sera and Plasma for Immunochemical Analysis to Detect Antibodies Against Infectious Pathogens (AbaCoV) Project” of DOST-PCHRD. Mr. Reneir John G. Tuason also participated in sample collection.

## SUPPLEMENTARY MATERIAL

Supplementary figures are hosted in figshare.

**Supplementary Figure 1**. Epitope map of the SARS-CoV-2 peptide library used for screening.

**Supplementary Figure 2**. LC-MS analysis confirming peptide identity and purity.

**Supplementary Figure 3**. Avidity calculation was facilitated by depolymerization verified by SDS-PAGE and indirect ELISA.

**Supplementary Figure 4**. ELISA optimization for lead candidate S559 with M212, N10, and E1 as comparator peptides.

**Supplementary Figure 5**. Participant flow diagram.

**Supplementary Figure 6**. Diagnostic accuracy was determined by indirect ELISA on clinical samples and biobanked controls using polymeric S559.

